# Posterior-anterior striatal reorganization after weight restoration in anorexia nervosa is associated with remission outcome: from mice to longitudinal human study

**DOI:** 10.64898/2026.10.23.26366116

**Authors:** Aynaz Adl Zarrabi, Arnaud Cachia, Virginie Tolle, Cindy Da Costa, Nicolas Lebrun, Clément Debacker, Sylvain Charron, Mallar M. Chakravarty, Stéphanie Daumas, Salah El Mestikawy, Catherine Oppenheim, Philip Gorwood, Philibert Duriez

**Affiliations:** Université Paris Cité, Institute of Psychiatry and Neuroscience of Paris (IPNP), INSERM U1266, Vulnerability to Psychiatric and Addictive Disorders, 75014 Paris, France; Université Paris Cité, Institute of Psychiatry and Neuroscience of Paris (IPNP), INSERM U1266, IMA-Brain, 75014 Paris, France; GHU-Paris Psychiatrie et Neurosciences, Hôpital Sainte-Anne, Clinique des maladies mentales et encéphales, F-75014 Paris, France; GHU-Paris Psychiatrie et Neurosciences, Hôpital Sainte-Anne, Imaging Department, F-75014 Paris, France; Douglas Mental Health University Institute, Department of Psychiatry, McGill University, Montreal, QC H4H 1R3, Canada; Cerebral Imaging Center, Douglas Mental Health University Institute, McGill University, Montreal, QC H4H 1R3, Canada; McGill University, Department of Biological and Biomedical Engineering, Montreal, QC H3A 2B4, Canada; Sorbonne Université, CNRS, Inserm, Centre of Neuroscience, Neuro-SU, Institut de Biologie Paris Seine, IBPS, F-75005 Paris, France; Université Paris Cité, Laboratory for the Psychology of Child Development and Education, CNRS UMR 8240, F-75005, Paris, France; Institut Universitaire de France (IUF)

**Keywords:** Anorexia nervosa, structural magnetic resonance imaging, remission, putamen, habits, ghrelin

## Abstract

Anorexia nervosa (AN) is a severe psychiatric disorder with high relapse rates despite successful weight restoration. Altered reward processing and an imbalance between goal-directed and habitual control within the striatum have been proposed to contribute to the persistence of maladaptive behaviours, but the striatal mechanisms underlying recovery and relapse remain unclear. Here, we combined cross-species structural MRI, including cross-sectional analyses in mice and longitudinal neuroimaging and clinical assessments in patients with AN, to investigate striatal alterations associated with recovery. Male mice with deletion of the vesicular acetylcholine transporter (VAChTcKO; n = 9), a model of maladaptive eating behaviour, showed reduced dorsolateral striatal volume compared with wild-type mice (n=9). At baseline, women with AN (n = 71) showed reduced striatal volumes compared with healthy controls (n = 30). Longitudinally, patients with AN (n = 41) and healthy controls (n = 28) were reassessed, revealing substantial recovery of striatal volumes following weight restoration in AN. Posterior-anterior putamen gradient changes differed according to subsequent remission status, with changes in the right putamen gradient predicting relapse. Striatal gradient changes were also associated with goal-directed and habitual behaviour during the slip-of-action task, response consistency during delay discounting, and circulating ghrelin and desacyl ghrelin concentrations. These findings identify longitudinal putamen reorganization as a candidate marker of vulnerability to relapse and link striatal plasticity, goal-directed and habitual control, reward processing, and metabolic processes in AN.

## Introduction

Anorexia nervosa (AN) is a severe psychiatric disorder characterized by persistent restriction of energy intake, intense fear of gaining weight, cognitive rigidity, and disturbances in body image perception^1,2^. AN affects approximately 1-4% of women^3^ and is associated with one of the highest mortality rates among psychiatric disorders^4^. Current treatment strategies primarily aim to restore body weight and alleviate eating disorders-related symptoms^5^. Despite successful weight restoration during treatment, relapse remains frequent, with nearly half of patients relapsing within the year following hospitalization^6–9^. Weight restoration does not necessarily indicate sustained clinical recovery, and the neurobiological changes associated with subsequent remission or relapse remain poorly understood^10^. Identifying neurobiological markers that distinguish sustained recovery from vulnerability to relapse therefore represents a major unmet clinical need^11^.

One proposed contributor to the persistence of AN is the development of maladaptive behaviours that become increasingly automatic and resistant to change. Repeated behaviours such as dietary restriction, excessive exercise and body checking may progressively become less sensitive to their consequences and more strongly driven by learned stimulus-response associations^12–14^. This has motivated the hypothesis that an imbalance between goal-directed and habitual control contributes to symptom persistence and relapse in AN^15–17^. However, evidence for increased habitual responding in AN remains mixed, suggesting that restrictive behaviours may reflect both persistent goal-directed pursuit of weight-related outcomes and increasingly automatic action patterns, rather than a simple shift from goal-directed to habitual control.^18,19^. These processes may be further shaped by altered reward and punishment sensitivity, as restrictive behaviours may be reinforced by anticipated weight-related outcomes and the avoidance of perceived adverse consequences of eating^20,21^.

These complementary behavioural systems are supported by partially segregated corticostriatal circuits involved in reward valuation, reinforcement learning and action selection^22^. Goal-directed actions remain sensitive to the expected value of their outcomes, whereas habitual actions become less dependent on outcome value and more strongly influenced by learned stimulus-response associations. Across species, rodent dorsomedial and dorsolateral striatal territories broadly correspond to associative and sensorimotor corticostriatal territories involving the human caudate and putamen, respectively, although these anatomical and functional correspondences are not one-to-one^23,24^. Striatal dopamine and acetylcholine signalling contribute to corticostriatal plasticity and to the balance between flexible, habitual and compulsive action patterns relevant to eating pathology^25,26^. Consistent with this framework, mice lacking the vesicular acetylcholine transporter (VAChT) in striatal cholinergic interneurons show impaired behavioural flexibility, enhanced habit formation and increased susceptibility to binge-like and anorexia-like behaviours^27–29^. Together, these findings implicate striatal cholinergic signalling in behavioural flexibility and habit formation, providing a rationale for examining striatal morphology across experimental and clinical contexts.

Human neuroimaging studies have implicated corticostriatal circuits in AN, particularly through alterations in functional activation and structural or functional connectivity^30,31^. Previous neuroimaging studies have also linked changes in anterior cingulate morphology and prefrontal activation to subsequent clinical outcomes in AN, but the prognostic relevance of spatially heterogeneous striatal reorganization remains largely unexplored^32–35^. Cross-sectional and longitudinal structural MRI studies have also demonstrated widespread gray matter alterations in AN, including changes in striatal morphology and volume, with substantial but often incomplete normalization following weight restoration^36,37^. However, conventional regional volumetric analyses provide limited information about whether structural changes are spatially homogeneous within the striatum or preferentially affect territories associated with distinct corticostriatal circuits. Characterizing such spatial heterogeneity may therefore offer additional insight into striatal reorganization during recovery.

The striatum exhibits spatial gradients in anatomical organization, functional specialization and corticostriatal connectivity^38,39^. These gradients encompass posterior sensorimotor territories associated with habitual action control and more anterior associative territories implicated in flexible, goal-directed behaviour^40,41^. Examining the spatial distribution of gray matter volume (GMV) along this axis may reveal heterogeneous patterns of structural change that are obscured by whole-region measurements^42,43^. Although morphometric gradients do not directly measure the functional balance between goal-directed and habitual systems, they may provide an anatomically informed description of changes across striatal territories. Whether these structural patterns reorganize during weight restoration in AN, distinguish subsequent remission outcomes, or relate to behavioural alterations remains unknown.

To examine the behavioural relevance of striatal reorganization, measures of action control and reward-related decision-making offer complementary perspectives. The Slip-of-Action Task (SOAT) assesses sensitivity to outcome devaluation, providing an experimental measure relevant to goal-directed and habitual action control^44–46^. Delay discounting task, by contrast, characterizes preferences between immediate and delayed rewards, providing a complementary measure of reward-related decision-making and cognitive control, both of which have been implicated in AN^47–51^. Beyond these behavioural processes, metabolic signals may influence reward-related decision-making. Ghrelin, an orexigenic hormone implicated in energy homeostasis and reward-related processes^52,53^, can modulate dopaminergic signalling^54^. Notably, previous work in AN has identified an altered association between circulating desacyl-ghrelin and delay discounting^55^, while desacyl-ghrelin has also been associated with food-cue-related caudate activation^56^. Together, these findings motivate exploratory examination of behavioural and metabolic correlates of striatal morphology, without presupposing a direct mechanistic relationship.

In this study, we combined two complementary cohorts, comprising a mouse model with disrupted striatal cholinergic signalling (VAChTcKO) and a human cohort of patients with AN and healthy controls, using cross-sectional structural MRI in mice and cross-sectional and longitudinal MRI in humans. We investigated whether (i) striatal GMV and its posterior-anterior spatial distribution change during weight restoration in AN, (ii) these structural changes are associated with subsequent sustained remission or relapse, and (iii) they relate to behavioural and circulating ghrelin measures, examining whether striatal reorganization during weight restoration was associated with subsequent clinical outcomes.

## Results

**Table 1:** Demographic information of participants included in this study.

|  | All participants |  |  |  | Participant status at V3 according to BMI |  |  |  |
| --- | --- | --- | --- | --- | --- | --- | --- | --- |
|  | AN | HC | Statistics | P | AN stable | AN relapsed | Statistics | P |
| Visit 1 | n=41 | n=28 | - | - | n=21 | n=18 | - | - |
| Age(Mean, SD) | 28.7 ± 10.9 | 26.8 ± 7.1 | U = 558.0 | 0.849 | 27.8 ± 10.6 | 30.2 ± 11.7 | U = 161.5 | 0.445 |
| Duration of illness(months) (Mean, SD) | 116.9 ± 121.2 | — | — | — | 114.0 ± 123.1 | 120.5 ± 128.1 | U = 165.5 | 0.517 |
| BMI (Mean, SD) | 14.4 ± 1.4 | 21.5 ± 1.9 | U = 0.0 | <0.001 | 14.8 ± 1.4 | 14.0 ± 1.3 | U = 267.0 | 0.029 |
| EDI-2 TOT (Mean, | 93.9 ± 41.7 | 20.2 ± 13.7 | U = 1088.5 | <0.001 | 99.8 ± 39.4 | 85.4 ± 45.4 | t = 1.05 | 0.299 |

|  |  |  |  |  |  |  |  |  |
| --- | --- | --- | --- | --- | --- | --- | --- | --- |
| SD) |  |  |  |  |  |  |  |  |
| HADS depression (Mean, SD) | 9.4 ± 4.1 | 2.8 ± 2.4 | U = 1073.0 | <0.001 | 9.1 ± 3.6 | 9.2 ± 4.7 | U = 200.5 | 0.755 |
| HADS anxiety (Mean, SD) | 13.0 ± 4.1 | 7.3 ± 3.0 | t = 6.69 | <0.001 | 13.4 ± 4.0 | 12.4 ± 4.4 | t = 0.73 | 0.469 |
| Visit 2 | n=41 | n=28 | - | - | n=21 | n=18 | - | - |
| BMI (Mean, SD) | 28.73 ± 10.89 | 26.79 ± 7.08 | U = 558.00 | 0.849 | 27.76 ± 10.63 | 30.17 ± 11.65 | U = 161.50 | 0.445 |
| EDI-2 TOT (Mean, SD) | 116.93 ± 121.16 | — | — | — | 114.05 ± 123.09 | 120.50 ± 128.06 | U = 165.50 | 0.517 |
| HADS depression (Mean, SD) | — | — | — | — | 20.31 ± 0.31 | 20.07 ± 0.36 | t = 2.26 | 0.03 |
| HADS anxiety (Mean, SD) | 73.90 ± 42.05 | 0.00 ± 0.00 | t = 11.25 | <0.001 | 73.86 ± 42.79 | 68.83 ± 41.40 | t = 0.37 | 0.713 |

### Demographic and clinical characteristics of the study cohorts

In the mouse cohort, male VAChTcKO mice (n = 9) and wild-type littermate controls (n = 9), aged 2-6 months, were included in the structural MRI analyses. The human cohort was drawn from the prospective REMANO protocol^57^, in which the baseline cross-sectional analyses included 71 women with anorexia nervosa (AN) and 30 healthy controls (HC), while longitudinal analyses included 41 AN patients and 28 HC with MRI data available at both baseline admission (V1) and discharge following weight restoration (V2) (Figure 2A). At baseline admission (V1), AN patients had a lower body mass index (BMI) than HC, while groups were comparable in age. AN patients also showed greater eating disorder symptom severity, anxiety and depressive symptoms, reflected by higher EDI-2^58^, HADS-Anxiety and HADS-Depression scores^59^ (all p < 0.001). Among patients with available follow-up outcome data approximately 6 months after hospitalization (V3), those who remained stable (n = 21) and those who relapsed (n = 18) did not differ at baseline in age, illness duration, eating disorder symptom severity, anxiety or depressive symptoms, although stable patients had slightly higher BMI at both admission and discharge.

### Altered dorsal striatal morphology in the VAChTcKO mouse model

VAChTcKO mice exhibit reduced cholinergic signaling within the dorsal striatum, comprising dorsomedial (DMS) and dorsolateral (DLS) territories (Figure 1A), together with an imbalance between goal-directed behavior and habits and increased vulnerability to anorexia-like self-starvation behavior in the activity-based anorexia (ABA) model^27,60^ . We first examined dorsal striatal morphology using cross-sectional structural MRI. VAChTcKO mice (n = 9) showed lower left DLS volume than wild-type controls (n = 9) (Welch t-test: 4.70 ± 0.43 vs 5.08 ± 0.25; t = 2.29, p = 0.039), although this difference did not survive FDR correction (p_FDR = 0.158). No group differences were observed in the right DLS (Welch t-test: WT, 5.35 ± 0.18; VAChTcKO, 5.13 ± 0.45; t = 1.35, p = 0.204), left DMS (WT, 5.16 ± 0.28; VAChTcKO, 4.99 ± 0.36; t = 1.13, p = 0.275), or right DMS (WT, 4.97 ± 0.21; VAChTcKO, 4.81 ± 0.37; t = 1.09, p = 0.298) (Figure 1B).Complementary deformation-based morphometry (DBM)^61^ showed a spatially consistent pattern of striatal tissue alteration, supporting the volumetric findings (Supplementary Figure S1).

**Figure 1:**
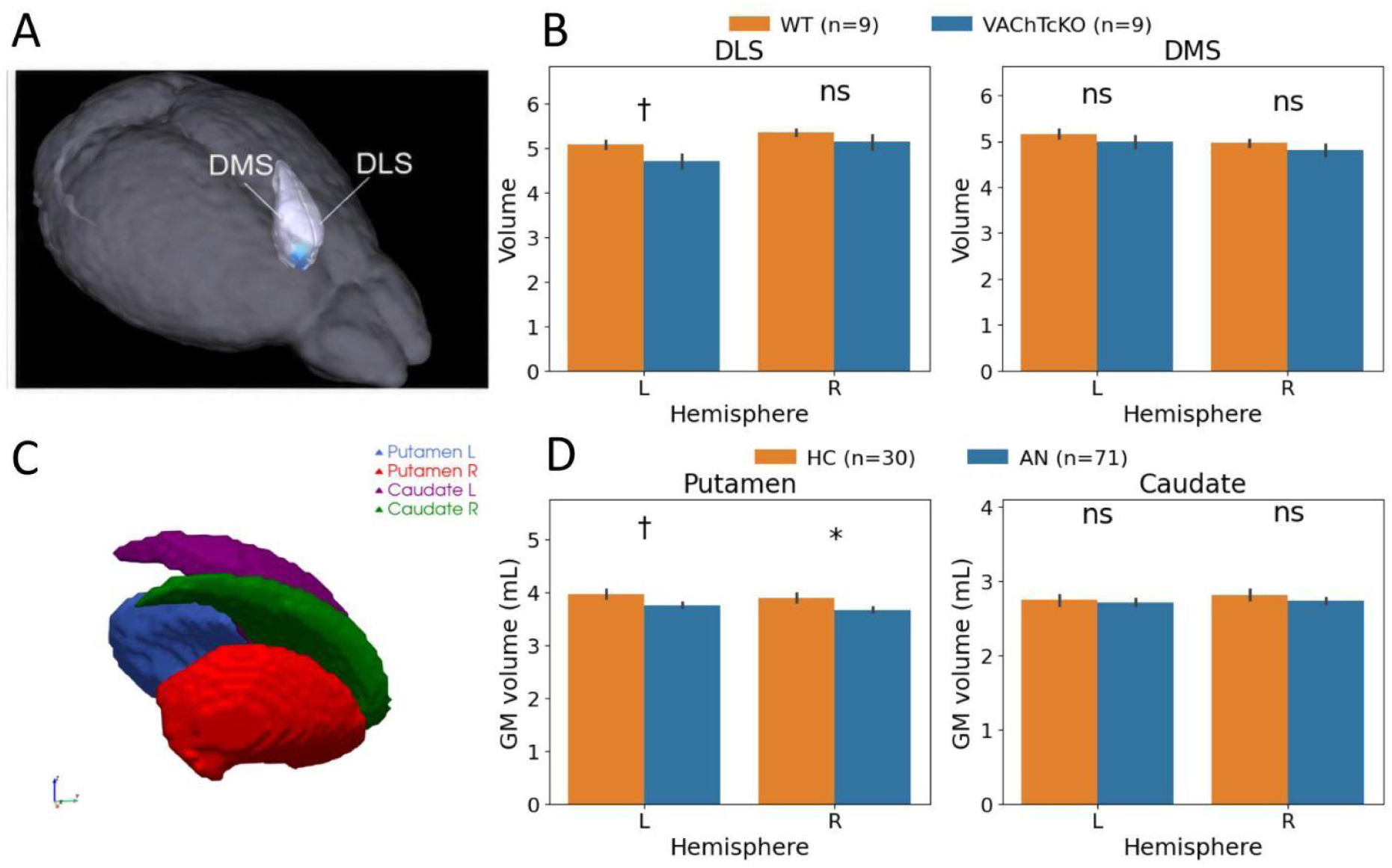
Morphological modification of the striatum of VAChTcKO mice that are vulnerable to anorexia-like phenotypes. Figure 1. (A) Three-dimensional anatomical localization of the dorsomedial and dorsolateral striatal territories in the mouse brain. (B) Regional striatal volumes in wild-type (WT) and VAChTcKO mice (n = 9/group). VAChTcKO mice showed lower left dorsolateral striatum (DLS) volume than WT controls (Welch’s t-test, p = 0.039), whereas right DLS and bilateral dorsomedial striatum (DMS) volumes did not differ significantly between groups. (C) Three-dimensional anatomical representation of the bilateral human putamen and caudate regions of interest. (D) Baseline gray matter volume (GMV) of the putamen and caudate in patients with anorexia nervosa (AN; n = 71) and healthy controls (HC; n = 30). GMV was lower in AN in the right putamen significatively (β = 0.160, pFDR = 0.032) and a nominally significant reduction in the left putamen (β = 0.142, p = 0.039, pFDR = 0.058), whereas caudate GMV did not differ significantly between groups. Error bars represent SEM. *p < 0.05; †*P* < 0.05 (uncorrected); ns, not significant.

### Parallel striatal abnormalities in anorexia nervosa

Focusing on the striatum, we examined the caudate and putamen (Figure 1C), the principal human homologues of the rodent DMS and DLS^23^. In our baseline cohort of 71 patients with anorexia nervosa (AN) and 30 healthy controls (HC), caudate GMV did not differ significantly between groups in either hemisphere (left: β = −0.023, p=0.69, pFDR = 0.737; right: β = 0.031, p=0.57, pFDR = 0.65). By contrast, putaminal GMV was lower in patients with AN than in healthy controls, with a significant difference in the right putamen (β = 0.160, pFDR = 0.032) and a nominally significant reduction in the left putamen (β = 0.142, p = 0.039, pFDR = 0.058) (Figure 1D). These findings suggest a similar pattern of preferential putaminal involvement to that observed in the VAChTcKO mouse model.<colcnt=1>

### Divergent trajectories of caudate and putamen in AN patients during weight restoration

Following the identification of baseline GMV alterations in the dorsal striatum (Table S1), we examined how these structures evolved during weight restoration. Longitudinal voxel-wise analyses^62^ in AN (n = 41) revealed widespread gray matter increases across most regions, whereas within the dorsal striatum, decreases were predominantly observed in the putamen (Table S2).

Region-of-interest (ROI) analyses^63^ further supported this dissociation between striatal subregions during weight restoration. Significant volume increases were observed in the bilateral caudate (left: β = 0.103, FDR-p = 2.95 × 10⁻¹⁸; right: β = 0.105, FDR-p = 2.42 × 10⁻¹⁵). By contrast, the left putamen exhibited a significant volume reduction (β = −0.129, FDR-p = 1.21 × 10⁻³), with a similar but non-significant trend in the right putamen (β = −0.053, FDR-p = 0.087) (Figure 2B). Healthy controls also underwent longitudinal MRI (n = 28), with no significant changes observed across global, voxel-wise, or ROI measures after correction for multiple comparisons.

**Figure 2:**
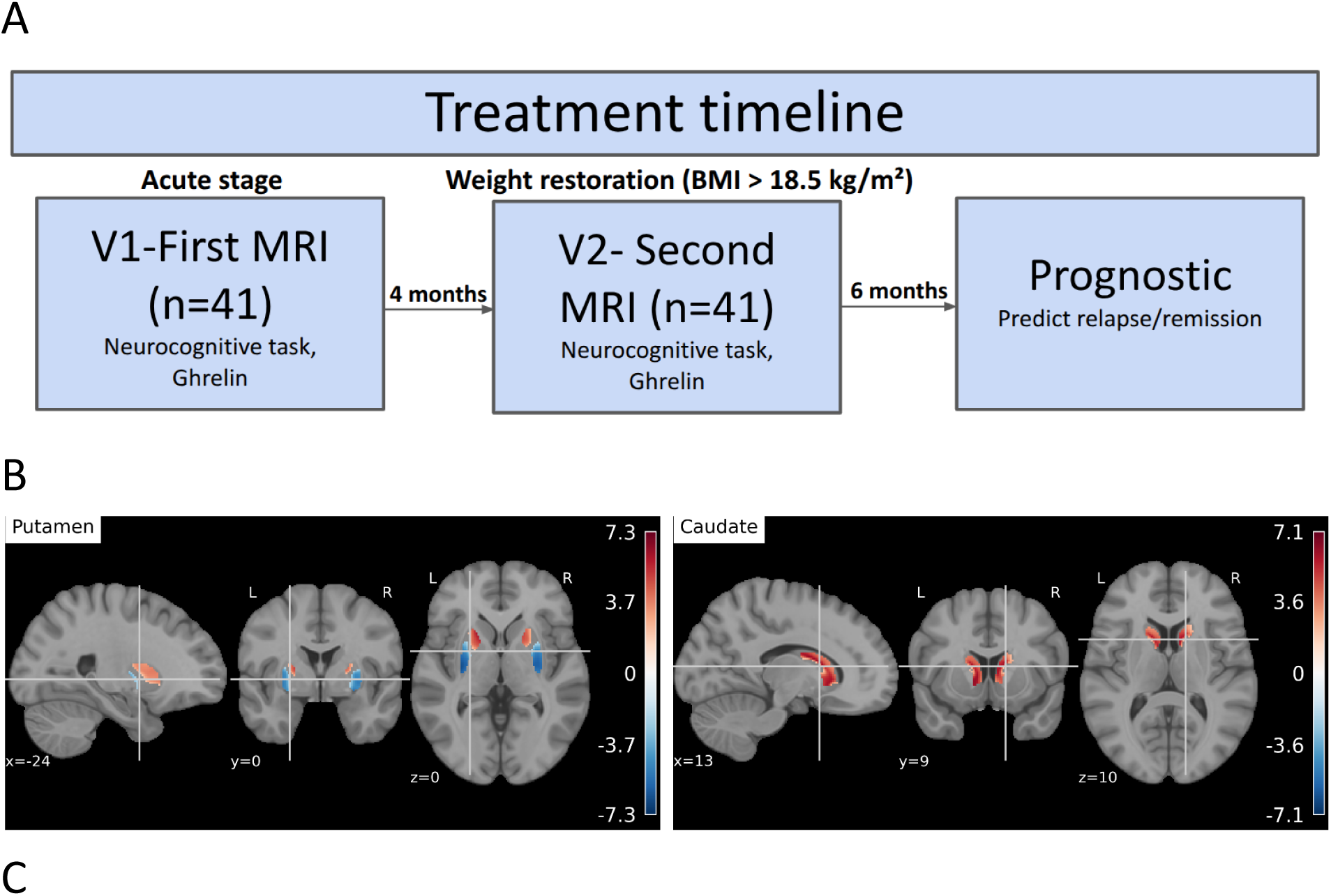
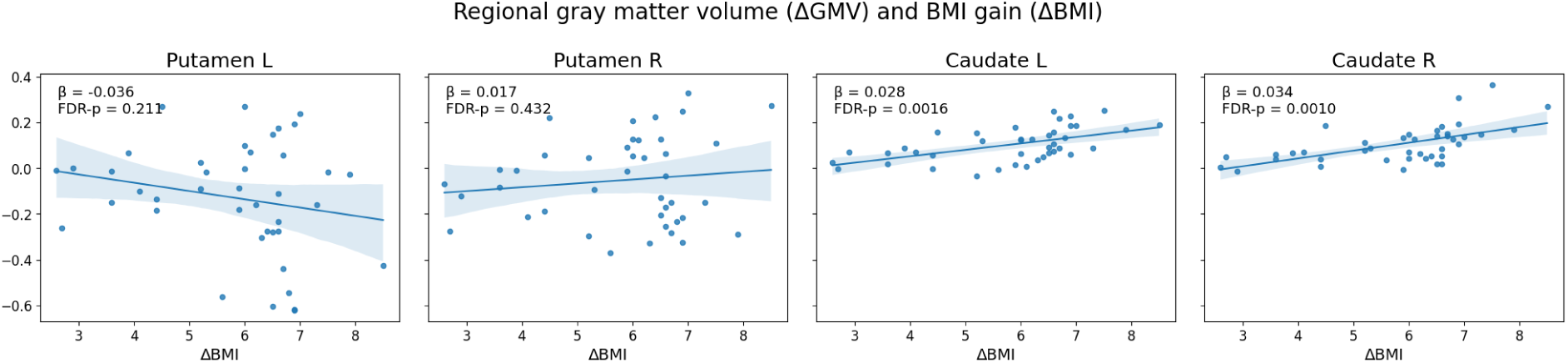
Longitudinal striatal volume changes following weight restoration in anorexia nervosa. Figure 2: (A) Schematic overview of the longitudinal study design, with MRI and neurocognitive assessment at baseline admission (V1), repeat assessment following weight restoration at discharge (V2), and clinical outcome assessment approximately 6 months later (V3). (B)Longitudinal voxel-wise gray matter changes from V1 to V2 in the putamen and caudate, regions implicated in habitual and goal-directed control, respectively, in patients with AN, displayed in MNI standard space, thresholded at FDR q < 0.05. (C) Associations between change in regional gray matter volume (ΔGMV) and BMI gain (ΔBMI) in the bilateral putamen and caudate. Greater BMI gain was associated with larger volume increases in the bilateral caudate, whereas putamen volume changes showed a negative association in the left hemisphere and a weak positive association in the right hemisphere, neither of which survived correction for multiple comparisons. Shaded areas indicate 95% confidence intervals.

Finally, to determine the extent to which these striatal changes reflected nutritional restoration, we examined their association with BMI gain. Greater BMI gain was significantly associated with larger volume increases in the bilateral caudate (left: β = 0.028, FDR-p = 0.0016; right: β = 0.034, FDR-p = 0.0010). By contrast, changes in putamen volume were not significantly associated with BMI gain after correction for multiple comparisons (left: β = −0.036, FDR-p = 0.211; right: β = 0.017, FDR-p = 0.432) (Figure 2C).These findings suggest that caudate volume changes are more closely coupled to weight restoration, whereas putaminal changes may reflect additional aspects of dorsal striatal remodeling not directly related to BMI gain.

### Longitudinal posterior-anterior striatal reorganization during weight restoration

Voxel-wise analyses revealed heterogeneous longitudinal changes within the putamen, contrasting with predominantly positive changes in the caudate (Figure 3A). This spatial heterogeneity suggested that whole-region measures may obscure changes occurring along the posterior-anterior axis.

**Figure 3:**
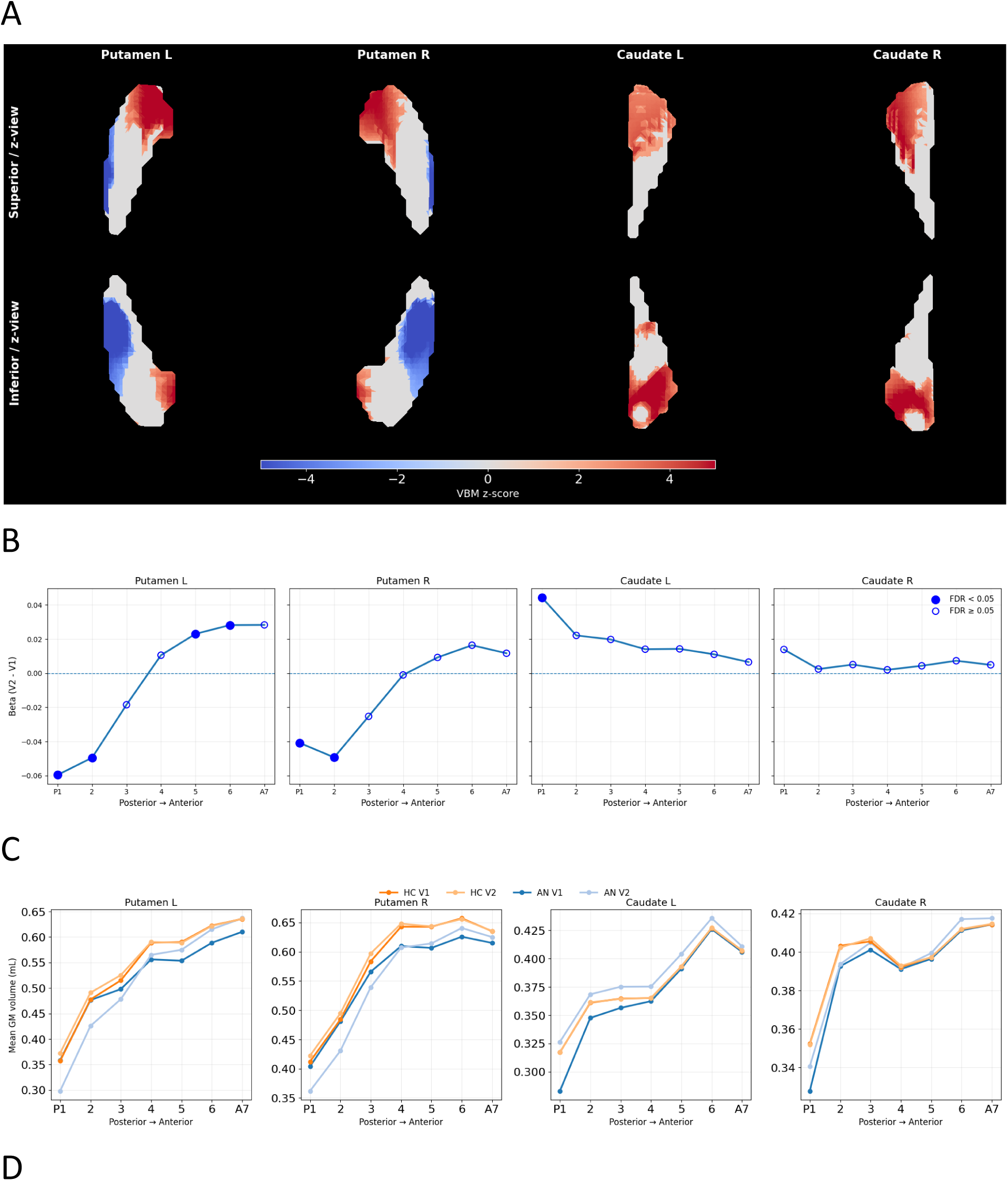
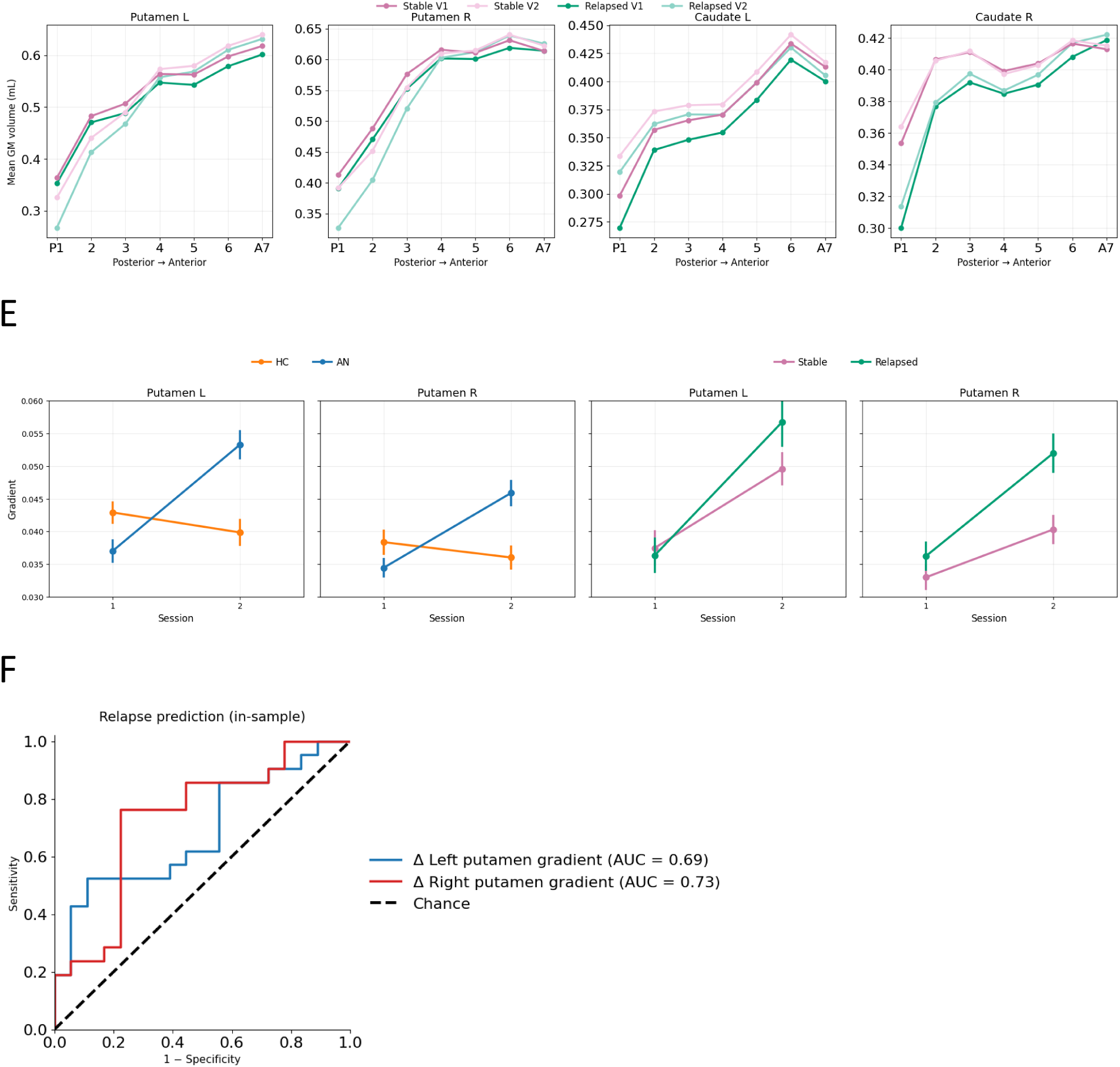
Longitudinal striatal gradient reorganization and prediction of relapse in anorexia nervosa. Figure 3: Longitudinal striatal gradient reorganization and prediction of relapse in anorexia nervosa (A) Three-dimensional surface projections of longitudinal voxel-based morphometry (VBM) changes from baseline admission (V1) to discharge following weight restoration (V2) within the bilateral putamen and caudate in MNI standard space. Red colors indicate relative gray matter increases and blue colors indicate decreases. (B) Segment-wise longitudinal gray matter volume changes (V2 − V1) across seven posterior-to-anterior segments of the bilateral putamen and caudate. Putaminal changes were negative in posterior and middle segments and became positive toward anterior segments, whereas caudate changes were predominantly positive across the posterior-anterior axis. Filled markers indicate FDR-corrected significance (P < 0.05), and open markers indicate nominal significance (P < 0.05). (C) Segment-wise gray matter volume profiles in healthy controls (HC) and patients with anorexia nervosa (AN) at V1 and V2, illustrating longitudinal reorganization of the posterior-anterior striatal profile. (D) Segment-wise gray matter volume profiles in AN patients according to subsequent remission status (stable versus relapsed), illustrating differences in longitudinal posterior-anterior organization between outcome groups. (E) Longitudinal changes in posterior-anterior putamen gradients in HC and AN and according to subsequent remission status. AN patients showed greater increases in bilateral putamen gradients than HC, while patients who subsequently relapsed showed a greater increase in the right putamen gradient than those with stable remission. Points represent mean gradient coefficients, and error bars indicate standard errors. (F) Receiver operating characteristic (ROC) curves illustrate the in-sample discrimination of stable versus relapsed patients using longitudinal changes in left and right putamen gradients, adjusted for age. The right putamen gradient showed the highest apparent discrimination (AUC = 0.73), followed by the left putamen gradient (AUC = 0.69).

**Figure 4.**
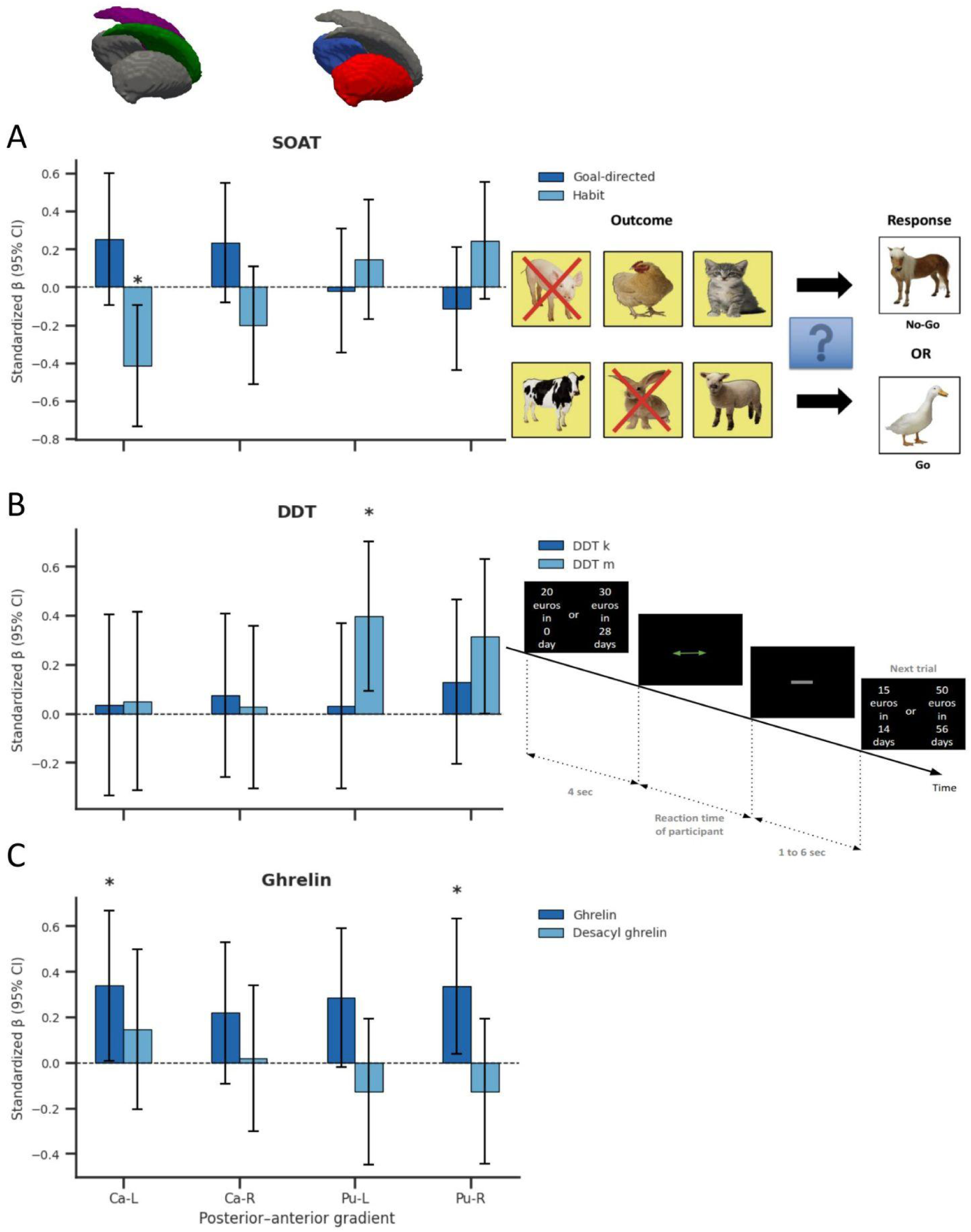
(A) Slip-of-Action Task schematic and associations between longitudinal changes in posterior-anterior. Figure 4: Neurocognitive, metabolic, and clinical correlates of striatal gradient reorganization. striatal gradients and changes in goal-directed and habitual responding (n = 40). Caudate gradients showed positive associations with goal-directed performance and negative associations with habitual responding, with the latter reaching nominal significance in the left caudate. In contrast, putamen gradients showed the opposite directional pattern, with negative associations with goal-directed performance and positive associations with habitual responding, although none reached statistical significance. (B) Delay-discounting task schematic and associations between longitudinal changes in striatal gradients and changes in the discount rate (k) and inverse temperature parameter (m) (n = 40). Associations with m were stronger than those with k, with the largest effect observed for the left putamen gradient. (C) Associations between longitudinal changes in striatal gradients and changes in circulating ghrelin and desacyl-ghrelin (n = 39). Changes in AG were positively associated with changes in the left caudate and right putamen gradients at nominal significance. In contrast, DAG showed opposite directional associations across striatal subregions, with positive coefficients in the caudate and negative coefficients in the putamen, although none reached statistical significance.

To characterize this spatial organization, we estimated posterior-anterior gradients from segment-wise GMV profiles along the caudate and putamen (see more in Methods). Longitudinal changes (V2-V1) revealed significant GMV reductions in posterior putaminal segments bilaterally (left: segments 1-2, FDR-p ≤ 0.0006; right: segments 1-3, FDR-p ≤ 0.042), whereas significant increases were observed in anterior segments of the left putamen (segments 5-7, FDR-p ≤ 0.031). Anterior segments of the right putamen showed positive but nonsignificant changes (segment 5: β = 0.0092, FDR-P = 0.333; segment 6: β = 0.0164, FDR-P = 0.110; segment 7: β = 0.0117, FDR-P = 0.323). In contrast, longitudinal changes in the caudate remained consistently positive across segments (Figure 3B). Together, these findings suggest spatially heterogeneous putaminal remodeling along the posterior-anterior axis during weight restoration in our cohort, rather than uniform volumetric recovery.

Consistent with the segment-wise GMV changes, posterior–anterior profiles revealed distinct longitudinal patterns in the caudate and putamen (Figure 3C). In the caudate, GMV profiles remained relatively similar between V1 and V2 within each group, whereas the putamen exhibited more pronounced longitudinal changes in spatial organization. To quantify these changes, we examined gradient coefficients derived from polynomial fits (see more in methods) to the segment-wise GMV profiles (Figure 3E, left). In patients with AN, bilateral putamen gradient coefficients increased significantly between V1 and V2 (left: β = +0.0163, FDR-P = 1.11 × 10⁻¹⁴; right: β = +0.0114, FDR-P = 6.10 × 10⁻¹³). Group × session analyses further revealed significantly greater longitudinal increases in putamen gradient coefficients in AN than in HC (left: β = −0.0194, FDR-P = 7.27 × 10⁻¹⁴; right: β = −0.0138, FDR-P = 9.88 × 10⁻¹¹). Together, these findings indicate more pronounced longitudinal reorganization of posterior–anterior putaminal GMV profiles in AN compared with HC.

### Posterior-anterior organization and remission outcome

We then investigated whether posterior-anterior striatal reorganization differed according to subsequent remission outcome. Gradient-based measures revealed differences between stable and relapsed patients (Figure 3D and E right), suggesting that spatially organized changes along the posterior-anterior axis may provide additional information beyond global volumetric measures. Specifically, relapsed patients showed a significantly greater increase in the right putamen gradient than patients with stable remission (β = −0.0084, FDR-P = 0.0278), indicating a more pronounced anterior shift of putaminal volume. A similar difference was nominally significant in the left putamen (β = −0.0083, P = 0.0479; FDR-P = 0.0958). No significant remission-group differences remained after FDR correction for the caudate gradients.

We further assessed whether longitudinal gradient changes (Δ gradients) were associated with subsequent remission outcome. Greater increases in the right putamen gradient were associated with lower odds of stable remission (standardized β = −0.98, OR per SD = 0.37, p = 0.020, FDR-p = 0.122; AUC = 0.73), with a similar association observed for the left putamen (standardized β = −0.80, OR per SD = 0.45, p = 0.052, FDR-p = 0.143; AUC = 0.69). Although neither association survived FDR correction, the direction of the effects was consistent with the longitudinal mixed-effects analyses, in which patients who subsequently relapsed exhibited greater increases in putamen gradients than those who achieved stable remission (Figure 3F). These findings should therefore be considered exploratory.

Complementary deformation-based morphometry (DBM) analyses supported the VBM findings by demonstrating heterogeneous longitudinal striatal tissue remodeling. ROI analyses based on DBM confirmed greater longitudinal expansion in the bilateral caudate and right putamen in AN than in healthy controls, consistent with the regional pattern observed using VBM. However, DBM measures were not significantly associated with remission outcome, in contrast to the posterior-anterior gradient measures, which showed significant or trend-level associations with clinical outcome (Supplementary Figure S2).

### Correlations with clinical outcomes Neurocognitive correlates

Goal-directed and habitual control were assessed using the Slip-of-Action Task (SOAT) through corresponding behavioural indices (see more in methods). At baseline, patients with AN showed lower goal-directed indices than healthy controls (51.34 ± 39.13 vs. 80.39 ± 27.22; FDR-P = 0.0017) and higher habitual responding (22.67 ± 24.73 vs. 10.56 ± 17.01; FDR-P = 0.0227) (Figure S4). Longitudinally (n = 40), greater increases in the left caudate posterior-anterior gradient were associated with higher goal-directed indices (β = 0.25, *P* = 0.1478) and lower habitual responding (β = −0.42, *P* = 0.0125; FDR-*P* = 0.0998). In contrast, putamen gradients showed an opposite directional pattern, with negative associations with goal-directed indices (left: β = −0.02, P = 0.9001; right: β = −0.11, P = 0.4780) and positive associations with habitual responding (right: β = 0.24, P = 0.1182). None of these longitudinal associations survived FDR correction.

Delay discounting was assessed by estimating two parameters: the discount rate (*k*), reflecting sensitivity to delayed rewards, and inverse temperature (*m*), reflecting response consistency^64^ (see more in methods). None of the DDT parameters showed significant longitudinal change from V1 to V2 (all P > 0.23) (Figure S3). Nevertheless, exploratory analyses (n = 40) suggested an association between longitudinal striatal reorganization and DDT performance, with greater increases in the left putamen posterior-anterior gradient associated with increases in m (β = 0.40, P = 0.0126; FDR-P = 0.101). A similar but weaker positive association was observed for the right putamen gradient (β = 0.31, P = 0.0508; FDR-P = 0.203), whereas associations involving the discount rate k were weaker overall. None of these associations survived FDR correction.

### Metabolic correlates

Ghrelin, a well-characterized marker of nutritional state, was assessed through its acylated (AG) and desacylated (DAG) forms. At baseline, both acyl- and desacyl-ghrelin concentrations were higher in AN than HC (AG: 4.22 vs. 3.76, FDR-*P* = 7.91 × 10⁻⁴; DAG: 3.38 vs. 2.66, *P* < 0.001). Both decreased significantly following weight restoration (AG: 4.20 to 3.85, n = 39, FDR-*P* = 2.2 × 10⁻⁵; DAG: *P* = 0.014, FDR-*P* = 0.024), consistent with changes in nutritional state^65^(Figure S5). Exploratory analyses (n = 39) showed that greater longitudinal increases in AG were associated with greater increases in the right putamen (β = 0.34, *P* = 0.0287; FDR-*P* = 0.179) and left caudate gradients (β = 0.34, *P* = 0.0459; FDR-*P* = 0.179). In contrast, changes in DAG showed weak positive associations with caudate gradients (left: β = 0.15; right: β = 0.02) and weak negative associations with putamen gradients (left and right: β = −0.13), none of which reached nominal significance (all *P* > 0.40). No associations survived FDR correction. The observed caudate association may be considered alongside previous human findings linking higher fasting DAG levels to greater food-cue-related activation in the caudate^56^.

Bars represent standardized regression coefficients (β), and error bars indicate 95% confidence intervals. Dashed horizontal lines indicate β = 0. Asterisks indicate nominally significant associations (P < 0.05); none survived false discovery rate (FDR) correction. All analyses were adjusted for age and total intracranial volume (TIV). Ca-L, left caudate; Ca-R, right caudate; Pu-L, left putamen; Pu-R, right putamen; DDT, delay-discounting task; SOAT, Slip-of-Action Task.

## Discussion

In the present study, In the present study, we investigated dorsal striatal organization in anorexia nervosa using a cross-species structural imaging framework, combining analyses in VAChTcKO mice with longitudinal neuroimaging in patients with AN and healthy controls. We characterized structural changes during weight restoration using conventional morphometric measures and posterior-anterior GMV gradients and examined their behavioural, endocrine and clinical associations. Our principal finding is that dorsal striatal changes during weight restoration are spatially heterogeneous, with longitudinal putamen gradient alterations associated with subsequent remission status.

The cross-species findings reveal partially corresponding patterns of structural alterations in sensorimotor striatal territories, despite the distinct biological and clinical contexts of the mouse model and human AN. In VAChTcKO mice, structural alterations were observed in the dorsolateral striatum, whereas the dorsomedial striatum remained relatively preserved. A comparable pattern emerged in humans, with more pronounced alterations in the putamen than in the caudate. Recent experimental evidence linking transitions toward habitual responding to changes in dorsolateral striatal activity^66^ provides a potential functional context for these anatomical observations. Together, these findings highlight sensorimotor striatal territories as a relevant focus for investigating structural alterations in AN, while the underlying mechanisms and extent of cross-species correspondence remain unresolved.

Previous longitudinal MRI studies have shown widespread gray matter reductions in anorexia nervosa with substantial recovery following weight restoration^67^. However, our findings reveal a more complex pattern within the dorsal striatum: whereas caudate GMV increased relatively consistently, the putamen exhibited posterior reductions alongside anterior increases. This spatial heterogeneity is compatible with the organization of the striatum along continuous anatomical and functional gradients^38,41^. Functional differentiation between posterior and anterior putaminal territories has also been demonstrated in Parkinson’s disease, although its underlying pathology differs fundamentally from AN^68^. Importantly, posterior-anterior gradients captured longitudinal spatial changes beyond those observed using conventional ROI-based measures, suggesting that spatially resolved morphometry may provide complementary information to whole-region volumetric analyses.

The clinical relevance of this spatial reorganization was further supported by its association with subsequent remission status. Patients who later relapsed exhibited greater increases in posterior-anterior putamen gradients during weight restoration than those who achieved sustained remission, particularly in the right hemisphere. Although exploratory logistic analyses suggested potential prognostic value, these associations did not survive FDR correction, and their discriminative performance was assessed in-sample. Together, these findings suggest that structural changes accompanying weight restoration may reflect processes associated with longer-term clinical outcomes, highlighting a potential distinction between nutritional restoration and sustained recovery.

The behavioural associations provide functional context for these structural changes. Established corticostriatal models suggest that the caudate preferentially contributes to flexible goal-directed behaviour, whereas the putamen is more strongly implicated in stimulus-response habits^33^.Exploratory analyses revealed opposite directional associations between caudate and putamen gradient changes and SOAT performance. Caudate changes showed positive associations with goal-directed and negative associations with habitual responding, whereas putaminal changes showed the reverse pattern. Exploratory delay-discounting analyses also revealed nominal associations between putamen gradient changes and response consistency, suggesting a possible relationship with reward-related decision-making processes^69^. Although these patterns are compatible with the proposed functional specialization of dorsal striatal subregions, none survived correction for multiple comparisons, and they do not establish a functional dissociation or imply that structural gradients directly measure behavioural control or reward processing.

The relationship between striatal gradients and ghrelin-related measures provides a potential metabolic context for dorsal striatal reorganization during weight restoration. The distinct patterns observed for acyl-ghrelin and desacyl-ghrelin suggest that different aspects of ghrelin signalling may be associated with striatal structural changes. Previous evidence implicating ghrelin in dopaminergic reward processing and corticostriatal plasticity^70,71^ as well as AN pathophysiology^72^, offers a possible biological framework for these findings. However, these associations did not survive correction for multiple comparisons, and further research is needed to determine whether ghrelin signalling contributes to striatal remodeling or merely covaries with nutritional recovery.

Collectively, these findings highlight the potential clinical relevance of spatially resolved dorsal striatal organization in AN. Together with the cross-species anatomical observations and exploratory behavioural and metabolic associations, they raise new questions about the distinct contributions of caudate and putamen remodeling to sustained recovery and relapse vulnerability, warranting further investigation in larger longitudinal cohorts.

## Limitations

Several limitations should be acknowledged. First, remission analyses were based on a relatively modest longitudinal sample, limiting statistical power for subgroup analyses. Second, the human cohort included only female patients, whereas the experimental mouse cohort consisted exclusively of males. This sex difference may influence the interpretation of cross-species comparisons and should be considered in future studies involving larger, sex-balanced cohorts to assess the generalizability of the findings. Third, although posterior-anterior gradient measures differentiated remission groups and showed the strongest associations with behavioural and metabolic measures, most behavioural and hormonal correlations did not survive correction for multiple comparisons and should therefore be considered exploratory. Fourth, only two MRI time points were available, preventing characterization of the temporal dynamics of striatal remodeling throughout recovery. Fifth, conventional VBM and DBM analyses were used to characterize regional volume changes and tissue deformation but were not predictive of clinical outcome. Finally, the predictive analyses were based exclusively on posterior-anterior gradient measures, which require validation in larger independent longitudinal cohorts before their clinical utility can be established.

## Supporting information

https://github.com/aynazadl/remano-MRI

## Data Availability

The individual-level data generated and analysed during this study are not publicly available due to participant confidentiality and ethical restrictions. Requests for access to de-identified data may be directed to the corresponding author and will be considered subject to approval by the study sponsor and applicable ethical and legal requirements.

## Acknowledgements

We thank Damian Justo for his work on the delay discounting task (DDT), Lucile Thelliez and Maliesse Lui for MRI data acquisition, Marie Fadigas for her contributions to the REMANO project, and Mathieu Favier, Marco A. M. Prado, Vania F. Prado, Jurgen Germann, Daniel Gallino, and Erika Vigneault for their contributions to the mouse study.

## Contributors

This research was funded by the Fondation Adrienne et Pierre Sommer, hosted by the Fondation de France (#00081239), and by the Agence Nationale de la Recherche (ANR; ‘RewAN’ project, grant no. ANR-19-CE37-0020). Additional funding was provided by the Fondation de France (grant nos. 00159297/WB-2024-55049), and by the Fondation Université Paris Cité through the “Sauver la vie 2020” programme to support MRI acquisitions and biological assays. Biological assays were also supported by a private donation from Mr J. R. The work additionally received funding from the Agence Nationale de la Recherche through the STRIDE 2026 project (grant number ANR-26-CE17-5975).

Competing interests: None declared.

Patient and public involvement

Patients and/or the public were not involved in the design, or conduct, or reporting or dissemination plans of this research.

Patient consent for publication

Not applicable.

Data availability

The MRI and clinical data supporting the findings of this study are not publicly available owing to privacy and ethical restrictions but are available from the corresponding author upon reasonable request.

## Methods

### Animals

Animal care, handling, and all experimental procedures were conducted in accordance with the guidelines of the Canadian Council on Animal Care and were approved by the Facility Animal Care Committee of the Douglas Research Centre (protocols 2008-5643 and 2014-7479) or the Animal Care Committee of the University of Western Ontario (protocol 2008-127). All procedures also complied with the National Institutes of Health Guide for the Care and Use of Laboratory Animals and were approved by the Duke University Animal Care and Use Committee.

The vesicular acetylcholine transporter conditional knockout (VAChTcKO) mouse line was generated as previously described (Favier et al. 2020)^27,73^. Briefly, VAChT^flox/flox^ mice were crossed with hemizygous D2-Cre mice to generate mice lacking VAChT selectively in DRD2-expressing neurons (VAChT^D2-Cre-flox/flox^). Littermate VAChT^flox/flox^ mice served as wild-type controls.

Mice were housed in groups of two to four per cage (except when individually housed for behavioural testing) under standard laboratory conditions (22 ± 2 °C; 12 h light/dark cycle, lights on from 07:00 to 19:00), with food (except during behavioural food restriction) and water available ad libitum. All efforts were made to minimize animal suffering and to reduce the number of animals used. Animals were randomly assigned to experimental groups, and investigators were blinded to genotype whenever possible during data collection and analysis.

### Participants

Participants were recruited at GHU Paris Psychiatrie et Neurosciences (Sainte-Anne Hospital, Paris) as neuroimaging part of the longitudinal REMANO study of anorexia nervosa (NCT04560517, Duriez et al., 2024)^57^. Female patients meeting DSM-5 criteria for anorexia nervosa underwent MRI, clinical evaluation, and neurocognitive assessment at baseline (V1). Approximately 4 months after admission, following weight restoration (BMI > 18.5), patients underwent repeat clinical and neurocognitive assessments at discharge (V2), with a subset completing a second MRI acquisition. Clinical outcome was assessed approximately 6 months after hospitalization (V3) based on BMI, allowing patients to be classified as having achieved sustained remission or relapse. Healthy age-matched female controls underwent the same baseline MRI, clinical, and neurocognitive assessments, with a subset completing a second MRI acquisition at follow-up. Participants with neurological disorders, MRI contraindications, pregnancy, or inability to provide informed consent were excluded.

### Clinical, behavioural and hormonal measures for the human study

Participants underwent a comprehensive clinical, behavioural, neuropsychological, and metabolic assessment. Clinical variables included BMI, duration of illness, and AN subtype. Eating-disorder psychopathology was assessed using the Eating Disorder Inventory-2 (EDI-2)^58^ (Table1), comprising eleven subscales (Drive for Thinness, Bulimia, Body Dissatisfaction, Ineffectiveness, Perfectionism, Interoceptive Awareness, Interpersonal Distrust, Maturity Fears, Asceticism, Impulse Regulation, and Social Insecurity). Anxiety and depressive symptoms were assessed using the Hospital Anxiety and Depression Scale (HADS)^59^, and obsessive-compulsive symptoms using the Yale-Brown Obsessive-Compulsive Scale (Y-BOCS)^74^ (Supplementary Table 3,4,5). Reward-based decision-making and goal-directed/habitual control were assessed using the DDT and SOAT, respectively, as detailed below. Circulating ghrelin concentrations were measured as peripheral metabolic markers implicated in appetite regulation and reward processing^52,75^.

### MRI acquisitions for animal study

For brain perfusion, mice were anaesthetized with ketamine (100 mg·kg⁻¹) and xylazine (10 mg·kg⁻¹) diluted in saline, perfused with 30 mL phosphate-buffered saline (PBS) containing heparin (1 μL·mL⁻¹) and 2 mM ProHance® at a flow rate of 1 mL·min⁻¹, followed by 30 mL of 4% paraformaldehyde (PFA) containing 2 mM ProHance® at the same flow rate. After dissection, brains were post-fixed overnight in 4% PFA with 2 mM ProHance® at 4°C, then transferred to PBS containing 0.02% sodium azide and 2 mM ProHance® until imaging. MRI data were acquired on a Bruker 7T 30-cm bore scanner with AVANCE electronics using a three-dimensional FLASH sequence (70 μm isotropic resolution).

### MRI acquisitions for human study

Human MRI data were acquired on a 3T Vantage Galan 3T/XGO system (Canon Medical Systems Corporation, Otawara, Japan) equipped with a 32-channel head coil at the Groupe Hospitalier Universitaire (GHU) Paris Psychiatrie & Neurosciences. Structural imaging consisted of a three-dimensional T1-weighted MPRAGE sequence (TR = 6.5 ms, TE = 3 ms, TI = 950 ms, flip angle = 9°, FOV = 256 × 256 mm², acquisition matrix = 256 × 256, 160 sagittal slices, slice thickness = 1 mm, shot interval = 2500 ms, voxel size = 1 × 1 × 1 mm³).

### MRI processing and morphometric analysis

MRI data were converted from DICOM to NIfTI format and organized according to the standard Brain Imaging Data Structure (BIDS) specification using HeuDiConv^76,77^. Structural 3D T1 images were processed using the Computational Anatomy Toolbox (CAT12, version 12.9) implemented in SPM12, running under MATLAB R2018b^63,78^. Preprocessing included denoising, bias-field correction, affine registration, probabilistic tissue segmentation into gray matter (GM), white matter (WM), and cerebrospinal fluid (CSF) using tissue probability maps, followed by adaptive maximum a posteriori segmentation with partial volume estimation^79,80^. Images were spatially normalized to Montreal Neurological Institute (MNI) standard space using Geodesic Shooting registration^81^.

Given the longitudinal study design, T1-weighted images acquired at baseline (V1) and follow-up (V2) were additionally processed using the CAT12 longitudinal pipeline. The neuroplasticity model was selected because MRI sessions were separated by approximately four months and relatively subtle structural changes were expected.

Structural MRI data were retained after successful CAT12 processing and visual confirmation of adequate segmentation, with no major artefacts, gross mis-segmentation, or incomplete brain coverage. CAT12 image-quality metrics, including IQR, NCR, ICR, and image-boundary measures, were additionally reviewed to identify scans requiring further visual inspection.

### Morphometric analysis

ROI-based analyses were performed using the default CAT12 probabilistic atlas (Neuromorphometrics) and unsmoothed modulated GM maps to preserve regional anatomical specificity. Total intracranial volume (TIV) was estimated as the sum of GM, WM, and CSF volumes in native space and included as a covariate in all analyses.

For voxel-wise analyses, modulated GM maps were smoothed with an 8-mm full-width at half-maximum (FWHM) Gaussian kernel. Significant clusters were anatomically localized using the Harvard-Oxford probabilistic atlas.

### Posterior-anterior gradient analysis

To investigate the observed spatial heterogeneity, reflected by a posterior-anterior gradient within the regions of interest, we adopted the approach proposed by Drori et al. (2022)^41^ for characterizing microstructural gradients of the human striatum, building on the broader framework of macroscale brain gradients described by Bernhardt et al. (2022)^39^. Inspired by their method, we subdivided the caudate and putamen into seven consecutive segments with the same number of voxels and the same distance along their main anatomical axis. For each structure, regions of interest were derived from the Harvard-Oxford atlas, and singular value decomposition (SVD)^82^ was used to identify the principal spatial axis, corresponding to the posterior-anterior direction. GMV was then extracted separately for each of the seven segments. To summarize the posterior-anterior gradient profile into a single metric, we fitted a polynomial function to the GMV values across the seven segments for each subject. The fitted polynomial provided an individual quantitative description of the posterior-anterior gray matter gradient, allowing us to quantify and compare the spatial distribution of GMV within the caudate and putamen between groups and across sessions.

### Deformation-based morphometry

To complement the voxel-based morphometry (VBM) analyses, deformation-based morphometry (DBM)^61^ was performed by CAT12 to quantify voxel-wise tissue expansion and contraction from the deformation fields generated during nonlinear spatial normalization. Unlike VBM, which relies on tissue segmentation, DBM directly measures local anatomical deformations by analysing the Jacobian determinant of the nonlinear deformation field, allowing the detection of regional expansion and contraction independent of tissue classification^83^. For both human and mouse MRI data, Jacobian determinant maps were computed from the nonlinear registration fields and used as voxel-wise measures of local volumetric change. For the mouse MRI data, Jacobian maps were smoothed with a 200-μm Gaussian kernel before statistical analysis^83–85^ (Figure S2, S3).

### The Delay Discounting Task (DDT)

The reward based decision making was assessed by Delay Discounting Task (DDT)^64^. Participants made repeated choices between a smaller reward available sooner (“Smaller Sooner”, SS) and a larger reward available after a longer delay (“Larger Later”, LL). These choices were used to estimate the temporal discount rate (k) and inverse temperature parameter (m). Lower k values indicate weaker discounting of delayed rewards, reflecting a greater relative valuation of larger delayed rewards. Higher m values indicate greater consistency in choosing the option with the higher estimated subjective value. Further details are provided in the Supplementary Methods.

### Slip-of-action Task (SOAT)

Goal-directed and habitual control were assessed using the Slip-of-Action Task (SOAT)^44,45^ as a part of instrumental learning task. During this task, specific outcomes were devalued, and participants were instructed to respond only to stimuli associated with still-valued outcomes while withholding responses to stimuli associated with devalued outcomes. Because previously learned stimulus-response associations remained unchanged, successful performance required the use of action-outcome knowledge. Responses to valued outcomes indexed goal-directed control, whereas responses to devalued outcomes (“slips of action”) indexed habitual responding.

### Blood sampling and ELISA Immuno-Assays

Blood was collected at each visit after an overnight fast on BD Vacutainer tubes treated with 15% EDTA and Aprotinin 250 HIU ^71^. After collection, blood was kept at 4°C on ice then centrifuged within 2h (1000 g for 10 min at 4°C). Plasma samples were aliquoted and one aliquot was immediately acidified with HCl (final concentration of 0.1N) since acidification is known to preserve ghrelin stability. Samples were then stored at -80°C until the assay.

Plasma levels of ghrelin and desacyl ghrelin were measured using specific EIA (ghrelin: catalog no. A05106; desacyl ghrelin: catalog no. A05119; Bertin Bioreagents, Montigny-le-Bretonneux, France). All used samples came from acidified aliquot. External quality control of the same human plasma was used in all assays to ensure inter-assay stability. Coefficients of variation for human acyl and desacyl ghrelin were between 8-9% and 10-16% for intra- and inter-assay respectively.

## Statistical analysis

Statistical analyses were performed in Python (version 3.12.3). ROI-based analyses were conducted using the Statsmodels library (version 0.14.6)^86^. For cross-sectional comparisons, ordinary least squares (OLS) regression models were fitted to assess group differences while adjusting for relevant covariates, including age and total intracranial volume (TIV)^87^. For longitudinal analyses, linear mixed-effects models (MixedLM) were used to account for repeated measurements within participants, with subject-specific random intercepts and fixed effects for group, session, and their interaction.

For voxel-wise analyses, group-level statistical analyses were performed using the general linear model (GLM) framework implemented in Nilearn (version 0.10.0)^88^. Multiple-comparison correction was performed using the false discovery rate (FDR) procedure with a significance threshold of q < 0.05^86^. Significant clusters were anatomically localized using the Harvard-Oxford probabilistic atlas to facilitate interpretation of voxel-wise findings.

Depending on data normality, group comparisons were performed using independent-samples t-tests or Mann-Whitney U tests, whereas within-subject comparisons were assessed using paired t-tests or Wilcoxon signed-rank tests, as appropriate. These analyses were performed using SciPy (version 1.17.0)^89^. FDR correction was applied to families of ROI-based statistical tests.

To evaluate the predictive value of neuroimaging biomarkers, logistic regression models were implemented using Scikit-learn (version 1.8.0)^90^. Model performance was assessed using the area under the receiver operating characteristic curve (AUC) together with classification accuracy.

Neuroimaging results were visualized using Nilearn (version 0.13.1)^88^, including statistical maps projected onto MNI standard templates and region-of-interest representations. Additional statistical plots and figures were generated using Matplotlib (version 3.10.8)^91^ and Seaborn (version 0.13.2)^92^.

## Notes

### Competing Interest Statement

The authors have declared no competing interest.

### Author Declarations

The Comite de Protection des Personnes Sud-Ouest et Outre-Mer II gave ethical approval for the REMANO study (approval number: 19.07.26.54412). All participants provided written informed consent prior to participation.

