## Supplementary material for "Posterior-anterior striatal reorganization after weight restoration in anorexia nervosa is associated with remission outcome: from mice to longitudinal human study": https://github.com/aynazadl/remano-MRI

**Supplementary materials**

Table of Contents


- Table S1. Whole brain tissue volumes (VBM) of participants included in this study
- Table S2.  ROI-based analysis of participants included in this study
- Table S3. Other clinical outcomes: AN vs HC at Visit 1
- Table S4. Other clinical outcomes: AN stable vs relapsed patients at Visit 1
- Table S5. Other clinical outcomes: AN stable vs relapsed patients at Visit 2
- Figure S1. Deformation-based morphometry, MRI detection of morphological alterations of the brain of VAChTcKO mice
- Figure S2. Deformation-based morphometry of longitudinal striatal changes
- Additional method: DDT
- Figure S3. Delay-discounting parameters across groups and weight restoration
- Figure S4. Slip-of-action parameters across groups and weight restoration
- Figure S5. Ghrelin measures across groups and weight restoration


**Table S1: Global GM/WM/CSF**

At baseline, patients with AN showed lower global gray matter volume (GMV) and higher cerebrospinal fluid (CSF) volume than healthy controls, with no difference in white matter volume (WMV). Following weight restoration, GMV increased ($\beta$ = 16.33 mL, p = $7.01 \times 10^{-12}$) and CSF decreased ($\beta$ = −32.46 mL, p = $1.47 \times 10^{-20}$), while WMV showed a modest reduction ($\beta$ = -4.84 mL, p=$1.44 \times 10^{-3}$. Healthy controls showed no significant longitudinal changes. Global cortical thickness also increased significantly between V1 and V2 across the whole cortex and both hemispheres.

| Table S1: Whole brain tissue volumes (VBM) of participants included in this study | | | | | | | | |
|---|---|---|---|---|---|---|---|---|
| | All participants | | | | Participants status at V3 by their BMI | | | |
| | AN | HC | Statistics | P | AN stable | AN relapsed | Statistics | P |
| Visit 1 | n=41 | n=28 | - | - | n=21 | n=18 | - | - |
| Gray matter (mL cm³) (Mean, SD) | 602.3 ± 39.7 | 637.1 ± 54.8 | t = -3.1 | 0.003 | 636.2 ± 38.7 | 634.6 ± 44.8 | t = 0.7 | 0.49 |
| White matter (mL cm³) (Mean, SD) | 491.6 ± 41.5 | 485.9 ± 49.8 | t = 0.5 | 0.608 | 483.6 ± 37.8 | 499.5 ± 51.2 | t = -0.4 | 0.712 |
| Cerebo-spinal fluid (mL cm³) (Mean, SD) | 299.3 ± 40.7 | 245.1 ± 29.4 | t = 6.1 | <0.001 | 258.1 ± 67.3 | 268.1 ± 38.3 | t = -1.1 | 0.291 |
| Total intracranial volume (TIV, mL cm³) (Mean, SD) | 1379.3 ± 97.0 | 1365.8 ± 104.9 | t = 0.6 | 0.583 | 1377.9 ± 94.8 | 1402.1 ± 116.4 | t = 0.2 | 0.858 |
| Global Cortical Thickness (CT, mm) (Mean, SD) | 2.4 ± 0.1 | 2.5 ± 0.1 | | | 2.4 ± 0.1 | 2.4 ± 0.1 | | |

| Table S1: Whole brain tissue volumes (VBM) of participants included in this study | | | | | | | | |
|---|---|---|---|---|---|---|---|---|
| Visit 2 | n=41 | n=28 | - | - | n=21 | n=18 | - | - |
| Gray matter (mL cm³) (Mean, SD) | 625.1 ± 41.1 | 640.6 ± 53.9 | t = -1.4 | 0.179 | 626.2 ± 33.1 | 631.2 ± 47.1 | t = 0.0 | 0.978 |
| White matter (mL cm³) (Mean, SD) | 491.7 ± 43.0 | 485.3 ± 50.7 | t = 0.6 | 0.571 | 489.7 ± 36.4 | 501.3 ± 48.9 | t = -0.5 | 0.643 |
| Cerebo-spinal fluid (mL cm³) (Mean, SD) | 270.5 ± 38.6 | 242.7 ± 24.6 | t = 3.7 | <0.001 | 269.4 ± 48.6 | 271.2 ± 27.3 | t = -0.2 | 0.854 |
| Total intracranial volume (TIV, mL cm³) (Mean, SD) | 1398.7 ± 96.9 | 1364.6 ± 103.7 | t = 1.4 | 0.168 | 1397.5 ± 90.5 | 1415.7 ± 103.9 | t = -0.2 | 0.813 |
| Global Cortical Thickness (CT, mm) (Mean, SD) | 2.4 ± 0.0 | 2.5 ± 0.1 | | | 2.4 ± 0.1 | 2.4 ± 0.1 | | |

| Table S2: ROI-based analysis of participants included in this study | | | | | | | | | | |
|---|---|---|---|---|---|---|---|---|---|---|
| Brain region | Side | x | y | z | Peak Z | ROI voxels | Significant voxels | % significant | % V2 > V1 | % V2 < V1 | Direction |
| NACC | L | -10.5 | 15 | -4.5 | 6.94 | 229 | 228 | 99.6 | 100 | 0 | V2 > V1 |
| NACC | R | 9 | 16.5 | -3 | 6.02 | 202 | 198 | 98 | 100 | 0 | V2 > V1 |
| caudate | L | -12 | 15 | 0 | 7.12 | 1175 | 953 | 81.1 | 100 | 0 | V2 > V1 |
| caudate | R | 10.5 | 1.5 | 16.5 | 6.47 | 1223 | 937 | 76.6 | 100 | 0 | V2 > V1 |
| putamen | L | -28.5 | -10.5 | -10.5 | -6.87 | 1897 | 1297 | 68.4 | 50.1 | 49.9 | V2 > V1 |
| putamen | R | 27 | -10.5 | -6 | -7.32 | 1867 | 1266 | 67.8 | 42.3 | 57.7 | V2 < V1 |
| Insula | whole | -36 | -12 | -3 | -6.7 | 6344 | 4400 | 69.4 | 67.9 | 32.1 | V2 > V1 |
| Amygdala | L | -27 | -9 | -13.5 | -6.77 | 727 | 495 | 68.1 | 30.1 | 69.9 | V2 < V1 |
| Amygdala | R | 27 | -7.5 | -12 | -6.42 | 842 | 568 | 67.5 | 24.8 | 75.2 | V2 < V1 |
| OFC | lateral | -40.5 | 24 | -9 | 6.36 | 7489 | 4457 | 59.5 | 93.5 | 6.5 | V2 > V1 |
| OFC | medial | 1.5 | 54 | -9 | 5.77 | 2302 | 1054 | 45.8 | 93.2 | 6.8 | V2 > V1 |
| ACC | Midline | 0 | -10.5 | 45 | 6.76 | 6122 | 2802 | 45.8 | 93.3 | 6.7 | V2 > V1 |

### Table S3 : other clinical outcomes AN vs HC Visit 1

| Variable | AN | HC | Statistics | P |
|---|---|---|---|---|
| n | 41 | 28 | - | - |
| HADS total | 22.4 ± 6.9 | 10.1 ± 4.8 | t = 8.8 | < 0.001 |

| | | | | |
|---|---|---|---|---|
| HADS anxiety | 13.0 ± 4.1 | 7.3 ± 3.0 | t = 6.7 | < 0.001 |
| HADS depression | 9.4 ± 4.1 | 2.8 ± 2.4 | U = 1073.0 | < 0.001 |
| EDI-2: Drive for thinness | 10.0 ± 7.2 | 0.4 ± 0.8 | U = 1078.0 | < 0.001 |
| EDI-2: Bulimia | 1.5 ± 4.0 | 0.5 ± 1.1 | U = 618.0 | 0.482 |
| EDI-2: Body dissatisfaction | 13.7 ± 7.0 | 2.4 ± 3.2 | U = 1081.0 | < 0.001 |
| EDI-2: Ineffectiveness | 13.5 ± 7.5 | 2.0 ± 2.6 | U = 1057.5 | < 0.001 |
| EDI-2: Perfectionism | 7.7 ± 4.3 | 4.7 ± 3.8 | U = 802.5 | 0.005 |
| EDI-2: Interoceptive awareness | 11.3 ± 7.2 | 1.4 ± 1.8 | U = 1066.5 | < 0.001 |
| EDI-2: Interpersonal distrust | 6.8 ± 4.1 | 1.6 ± 2.5 | U = 1005.5 | < 0.001 |
| EDI-2: Maturity fears | 6.6 ± 6.2 | 2.3 ± 2.6 | U = 835.5 | 0.001 |
| EDI-2: Asceticism | 8.8 ± 5.1 | 1.5 ± 1.5 | U = 1051.0 | < 0.001 |
| EDI-2: Impulse regulation | 4.8 ± 3.9 | 1.2 ± 1.7 | U = 930.5 | < 0.001 |
| EDI-2: Social insecurity | 9.3 ± 4.9 | 2.2 ± 2.6 | U = 1030.0 | < 0.001 |
| YBC-EDS: Obsessions | 9.4 ± 5.2 | 3.0 ± 3.2 | U = 962.0 | < 0.001 |
| YBC-EDS: Compulsions | 9.2 ± 5.1 | 2.9 ± 3.7 | U = 947.0 | < 0.001 |

### Table S4 : other clinical outcomes AN stable vs relapsed patients Visit 1

| Variable | AN stable | AN relapsed | Statistics | P |
|---|---|---|---|---|
| n | 21 | 18 | - | - |
| EDI-2: Drive for thinness | 10.9 ± 7.6 | 8.7 ± 7.1 | U = 222.0 | 0.359 |
| EDI-2: Bulimia | 1.6 ± 4.6 | 1.2 ± 3.6 | U = 226.0 | 0.164 |
| EDI-2: Body dissatisfaction | 14.9 ± 7.1 | 12.2 ± 7.0 | t = 1.2 | 0.253 |
| EDI-2: Ineffectiveness | 14.6 ± 6.9 | 12.1 ± 8.2 | t = 1.0 | 0.306 |
| EDI-2: Perfectionism | 8.3 ± 4.6 | 7.4 ± 4.1 | t = 0.6 | 0.526 |
| EDI-2: Interoceptive awareness | 11.6 ± 7.4 | 10.8 ± 7.4 | U = 203.0 | 0.703 |
| EDI-2: Interpersonal distrust | 6.9 ± 4.0 | 6.3 ± 4.5 | U = 206.0 | 0.641 |
| EDI-2: Maturity fears | 6.6 ± 5.6 | 6.7 ± 7.1 | U = 200.5 | 0.756 |
| EDI-2: Asceticism | 9.4 ± 5.4 | 7.5 ± 4.3 | t = 1.2 | 0.227 |
| EDI-2: Impulse regulation | 5.1 ± 4.2 | 4.2 ± 3.8 | U = 214.0 | 0.488 |
| EDI-2: Social insecurity | 10.0 ± 4.6 | 8.3 ± 5.6 | t = 1.1 | 0.295 |

| | | | | |
|---|---|---|---|---|
| YBC-EDS: Obsessions | 9.3 ± 5.6 | 9.9 ± 4.7 | t = -0.4 | 0.718 |
| YBC-EDS: Compulsions | 8.7 ± 4.8 | 10.1 ± 5.6 | t = -0.8 | 0.409 |

**Table S5 : other clinical outcomes AN stable vs relapsed patients Visit 2**

| Variable | AN stable | AN relapsed | Statistics | P |
|---|---|---|---|---|
| n | 21 | 18 | - | - |
| EDI-2: Drive for thinness | 8.8 ± 7.2 | 6.6 ± 7.3 | U = 220.5 | 0.379 |
| EDI-2: Bulimia | 1.1 ± 4.6 | 0.0 ± 0.0 | U = 207.0 | 0.197 |
| EDI-2: Body dissatisfaction | 12.2 ± 9.4 | 11.9 ± 10.0 | U = 191.5 | 0.955 |
| EDI-2: Ineffectiveness | 9.3 ± 6.6 | 9.0 ± 8.4 | U = 207.0 | 0.621 |
| EDI-2: Perfectionism | 6.8 ± 5.3 | 6.9 ± 4.4 | t = -0.1 | 0.908 |
| EDI-2: Interoceptive awareness | 6.8 ± 6.1 | 8.4 ± 6.0 | U = 158.0 | 0.389 |
| EDI-2: Interpersonal distrust | 5.0 ± 4.0 | 6.1 ± 3.9 | t = -0.9 | 0.387 |
| EDI-2: Maturity fears | 5.5 ± 6.0 | 5.9 ± 6.7 | U = 186.5 | 0.955 |
| EDI-2: Asceticism | 6.6 ± 4.6 | 4.6 ± 3.3 | U = 235.0 | 0.192 |
| EDI-2: Impulse regulation | 5.0 ± 5.5 | 2.2 ± 3.2 | U = 240.5 | 0.138 |
| EDI-2: Social insecurity | 6.9 ± 4.4 | 7.2 ± 5.2 | t = -0.2 | 0.865 |
| YBC-EDS: Obsessions | 7.1 ± 4.7 | 7.6 ± 4.0 | t = -0.4 | 0.715 |
| YBC-EDS: Compulsions | 6.3 ± 4.5 | 6.3 ± 5.0 | U = 184.0 | 0.898 |

**Figure S1**

DBM complemented the VBM findings by characterizing the local deformation patterns underlying the observed volumetric changes. Neuroanatomical differences between VAChTcKO mice and WT mice throughout the brain on coronal slices. Local volume increases in the mutant mice compared to WT are shown in red, local volume decreases are in blue. The t-thresholds are chosen to correspond with $p_{(uncorrected)}<0.05$ ($|t=2.09|$) and $p_{(uncorrected)}<0.001$ ($|t=3.68|$). The statistical maps are overlaid onto the group average. Positions of the slices, from caudal to rostral, are shown on the sagittal brain outline.

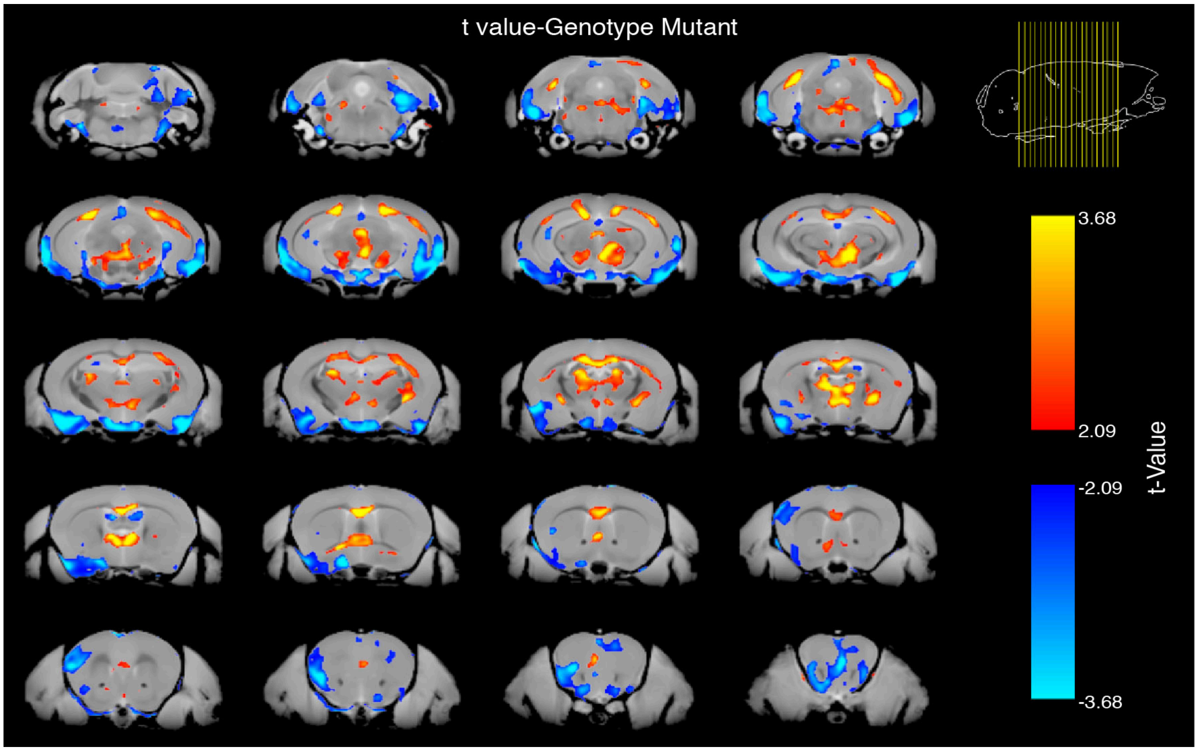


**Figure S2**

ROI-based analyses confirmed significantly greater longitudinal expansion in AN than in controls in the right caudate (β = 0.0208, FDR = $3.1 \times 10^{-7}$), left caudate (β = 0.0160, FDR = $2.8 \times 10^{-5}$), and right putamen (β = 0.0165, FDR = 0.021). Within the caudate, deformation was spatially heterogeneous, with widespread expansion in the anterior slices (85.3-93.4% of voxels showing V2 > V1) but substantial contraction in the posterior slices (38.8-48.7% of voxels showing V2 < V1). In contrast, the putamen showed predominantly expansive changes across the anterior-posterior axis, with 86.0-99.3% of voxels expanding between V1 and V2. Although these ROI-level DBM measures captured longitudinal tissue deformation, they did not significantly predict remission outcome, suggesting that the spatial organization of deformation, rather than its overall magnitude, may provide greater prognostic value. These findings indicate that structural recovery is not spatially uniform within the striatum and demonstrate that DBM provides complementary information on local tissue deformation beyond the regional volumetric changes identified by VBM.

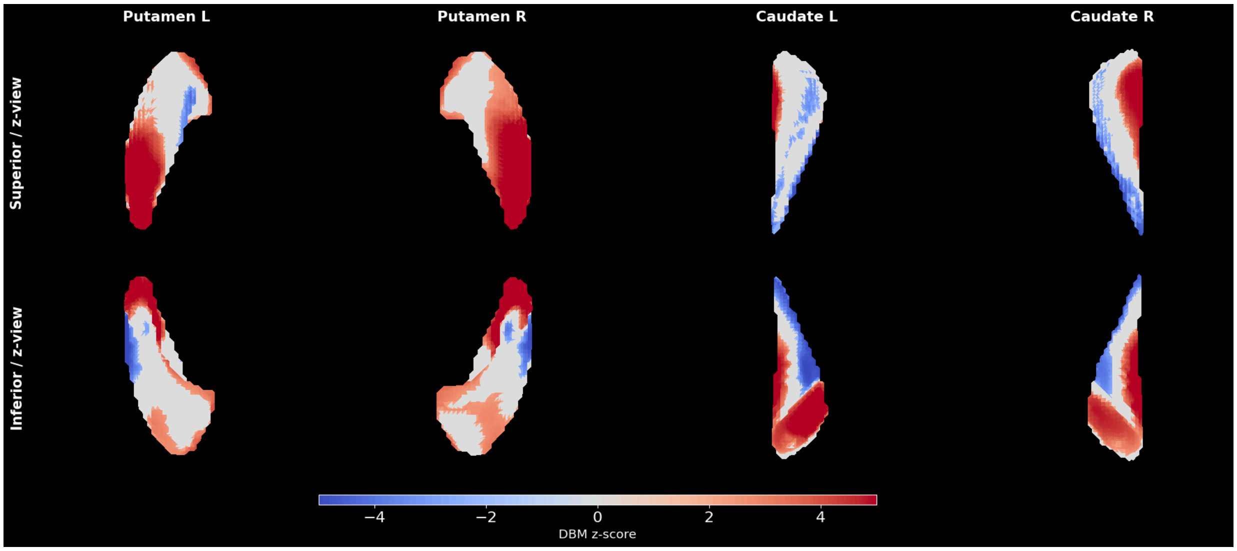


**Additional method : DDT**

According to the hyperbolic model describing the discounting function, the subjective value of a reward is described by the equation: $SV = \frac{A}{1+ K*D}$

where $SV$ refers to the subjective value of a reward with an object value $A$, available after a delay $D$. The coefficient $k$ is the discount rate which describes the individual differences regarding sensitivity to delay. $A$ smaller discount rate $k$ translates into a less steep discounting curve, meaning a higher perceived value of a given reward after a certain delay and consequently higher tolerance to waiting. Therefore, subjects with smaller k coefficients show a higher propensity to choose a larger and later reward.

The hyperbolic model will be used to determine the individual coefficient $k$ values for each participant. $LL = SS * (1 + K * D)$

The DDT will be carried out and serve to determine the individual coefficient $k$ to be identified following a staircase procedure. The $SS$ option is fixed on the amount of 20.00 euros and available the same day $d = 0$ and the $LL$ options will be generated according to the function:
The value of $k$ is initially set on 0.02 and will be progressively adjusted according to the participant's choices. After completion of the training phase, the individual $k$ will be identified using a grid search procedure, considering the choices of the participant throughout the trials. The rationality hypothesis yields that individuals will be more likely to choose the option with the highest subjective value. To take into account inconsistencies in subject choices, when estimating k, we use a softmax function, which introduces an additional parameter: the inverse temperature (m). Thus the probability of choosing the SS option ($P_{SS}$) depends on the subjective value of the $SS$ ($V_{SS}$) and the subjective value of the $LL(V_{ll})$, and is described according to the equation:

$$P_{SS} = \frac{e^{mV_{ss}}}{e^{mV_{ss}} + e^{mV_{ll}}}$$

with m the inverse temperature, a free parameter that captures the response noise i.e. the inconsistencies in the participant's choices. When m is equal to 0, $P_{SS}$ is equal to 0.5 ; the higher m, the more likely the subject will choose the option with the highest subjective value.

**Behavioural, metabolic associations**

**Figure S3**

At baseline (V1), goal-directed performance in SOAT task, differed significantly between patients with AN and healthy controls (AN: n = 40; HC: n = 28; U = 294.5, P < 0.001). Goal-directed performance did not change significantly following weight restoration from V1 to V2 (n = 40; W = 309.0, P = 0.258). Habitual responding also differed significantly between AN patients and healthy controls at baseline (AN: n = 40; HC: n = 28; U = 796.5, P = 0.003), but showed no significant longitudinal change following weight restoration (n = 40; W = 348.0, P = 0.958).

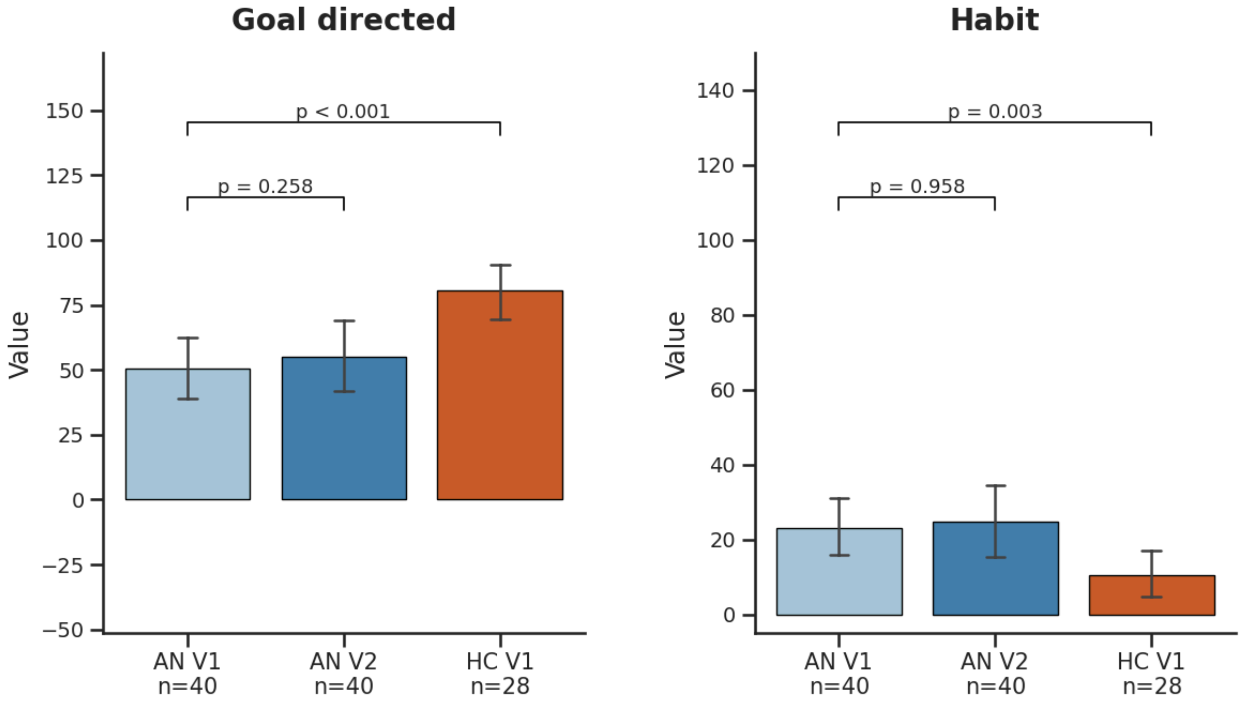


**Figure S4**

Delay-discounting parameters did not differ significantly between patients with AN and healthy controls at baseline and did not change significantly following weight restoration. For the discounting parameter k, there was no difference between AN at V1 and HC at V1 (U = 597, p = 0.649) and no significant change from V1 to V2 in AN (W = 321, p = 0.237). Similarly, the response-consistency parameter m did not differ between AN at V1 and HC at V1 (U = 536, p = 0.769) and did not change significantly from V1 to V2 (W = 222, p = 0.432).

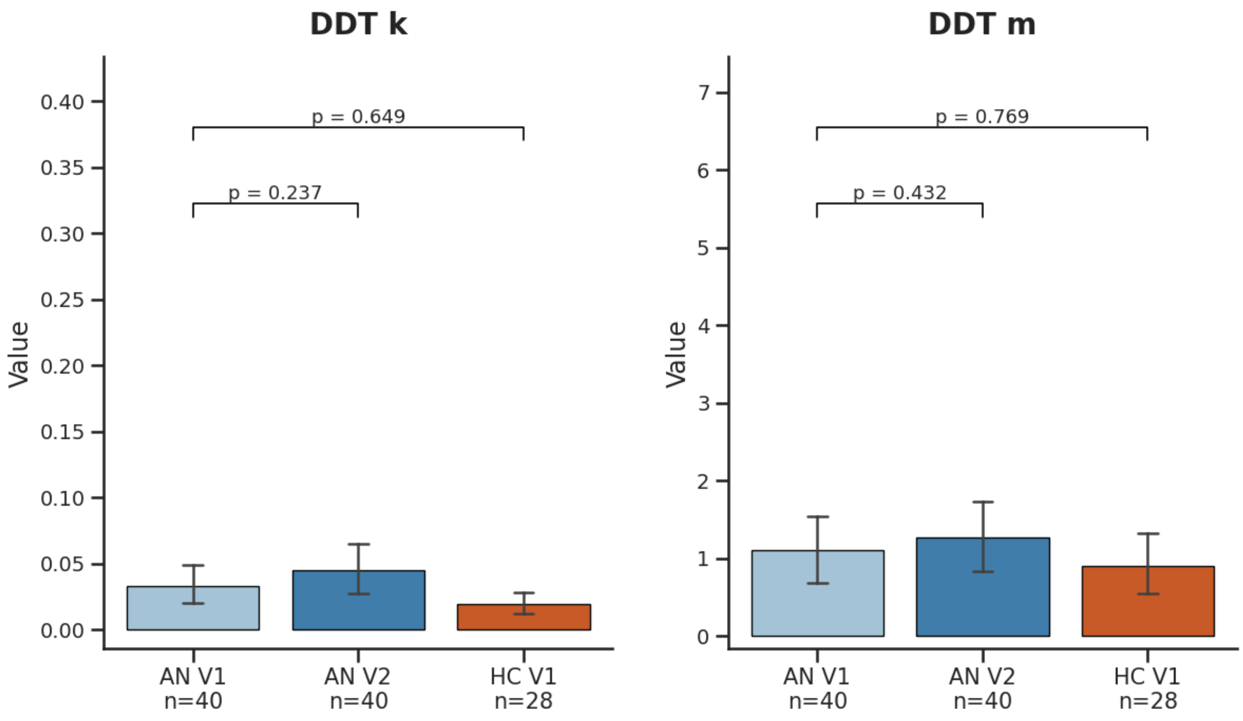


**Figure S5**

At baseline (V1), patients with AN had higher plasma ghrelin concentrations than healthy controls (AN: n = 40, mean log-transformed value = 4.22; HC: n = 28, mean = 3.76; $p < 0.001$). AG concentrations decreased significantly following weight restoration (AN V1: n = 39, mean = 4.20; AN V2: n = 39, mean = 3.85; $p < 0.001$; FDR-P = $5.5 \times 10^{-5}$). Desacyl ghrelin (DAG) concentrations were also higher at baseline in AN than in HC (AN: n = 40, mean = 3.38; HC: n = 28, mean = 2.66; $p < 0.001$) and decreased significantly from V1 to V2 (n = 39; $p = 0.014$; FDR-P = 0.024).

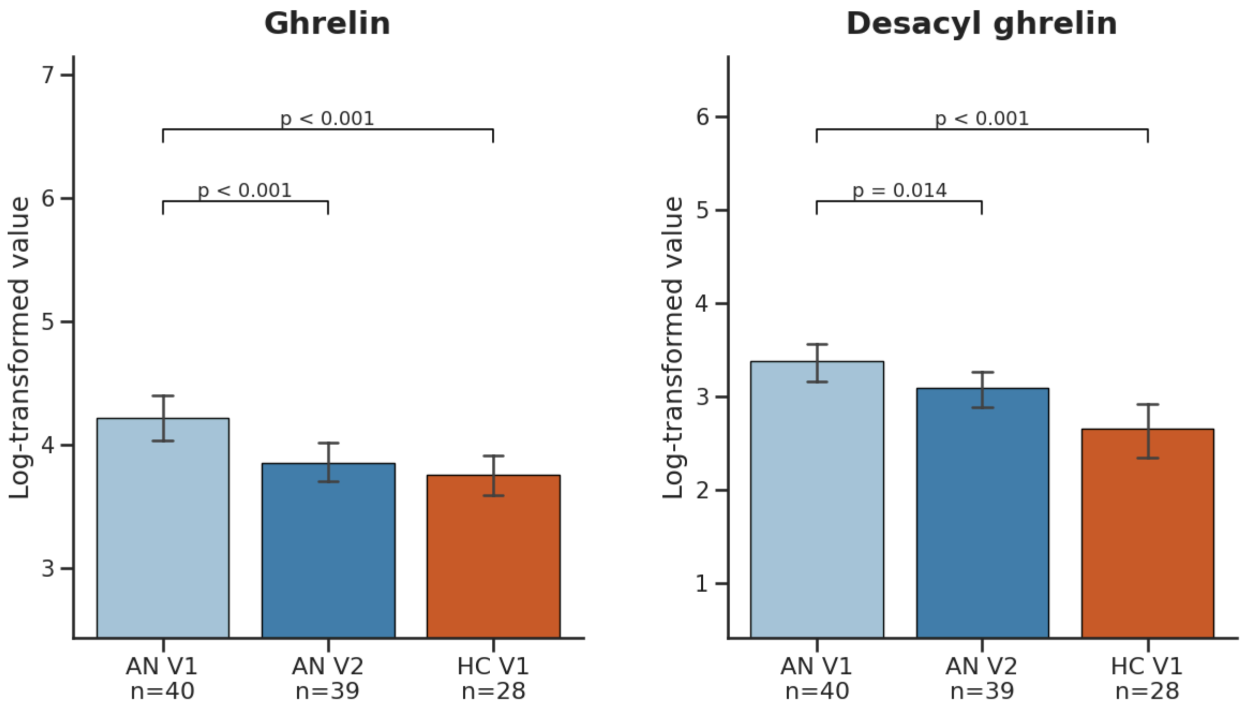
